# A qualitative analysis of community health worker perspectives on the implementation of the preconception and pregnancy phases of the *Bukhali* randomised controlled trial

**DOI:** 10.1101/2023.10.17.23297175

**Authors:** Larske M Soepnel, Shane A Norris, Khuthala Mabetha, Molebogeng Motlhatlhedi, Nokuthula Nkosi, Stephen Lye, Catherine E Draper

## Abstract

Community health workers (CHWs) play an important role in health systems in low– and middle-income countries, including South Africa. *Bukhali* is a CHW-delivered intervention as part of a randomised controlled trial, to improve the health trajectories of young women in Soweto, South Africa. This study aimed to qualitatively explore factors influencing implementation of the preconception and pregnancy phases of *Bukhali*, from the perspective of the CHWs (Health Helpers, HHs) delivering the intervention. As part of the *Bukhali* trial process evaluation, three focus group discussions were conducted with the 13 HHs employed by the trial. A thematic approach was used to analyse the data, drawing on elements of a reflexive thematic and codebook approach. The following six themes were developed, representing factors impacting implementation of the HH roles: interaction with the existing public healthcare sector; participant perceptions of health; health literacy and language barriers; participants’ socioeconomic constraints; family, partner, and community views of trial components; and the HH-participant relationship. HHs reported uses of several trial-based tools to overcome implementation challenges, increasing their ability to implement their roles as planned. The relationship of trust between the HH and participants seemed to function as one important mechanism for impact. The findings supported a number of adaptations to the implementation of *Bukhali*, such as intensified trial-based follow-up of referrals that do not receive management at clinics, continued HH training and community engagement parallel to trial implementation, with an increased emphasis on health-related stigma and education.

HH perspectives on intervention implementation highlighted adaptations relevant to the *Bukhali* intervention, other CHW-delivered preconception and pregnancy trials, and ‘real-world’ CHW roles, across three broad strategic areas: navigating and bridging healthcare systems, adaptability to individual participant needs, and navigating stigma around disease.

## 1. Introduction

Community health workers (CHWs) play an important and growing role in health systems in low– and middle-income countries, due to their potential to improve health at a community level while shifting tasks away from often overburdened formal healthcare sectors (1–3). As a cost-effective link between communities and primary healthcare services, CHWs also have the potential to strengthen health in contexts when regular access of healthcare services is otherwise rare, such as during the preconception period (4,5).

In South Africa, CHWs provide services, such as health screening, education, and advocacy, for various conditions, such as HIV, maternal and child health-related care, TB, and non-communicable diseases (NCDs) (2,6,7). However, previous research has identified challenges with implementing CHW programmes within the public health sector, including resource limitations, limited supervisor availability, low wages, and safety concerns (2). Particularly when contending with such contextual factors, insights are needed into the ways in which preconception and pregnancy intervention strategies with CHWs can strengthen health systems, and why they do or do not work.

### 1.1 CHWs in the *Bukhali* intervention: Health Helpers

*Bukhali* is a randomised controlled trial in South Africa, forming part of the Healthy Life Trajectories Initiative (HeLTI), a research initiative in collaboration with the World Health Organisation (WHO) with additional trials in India, Canada, and China. *Bukhali* is a complex, continuum of care intervention for 18-28 year old women, delivered by community health workers known as Health Helpers (HHs). It aims to improve women’s mental health, physical health, and nutrition to establish healthier trajectories for them and their future children, through four phases: preconception, pregnancy, infancy, and early childhood (up to five years), to offset obesity and NCD risk in offspring and improve development (8). *Bukhali* was developed with the goal of functioning within and complementing the realities of the South African public healthcare sector, and within Soweto, an urban, predominantly low-income setting in Johannesburg. Young women living in Soweto have been found to face a number of challenges in terms of their physical and mental health (9,10). In order to maximise the applicability and relevance of the intervention, a pragmatic approach has been adopted to the trial design (10,11), allowing for the incorporation of new learnings as the trial is implemented, and for adaptability in the face of challenges inherent to the trial context (8).

Due to its multi-phase nature, the emphasis of the *Bukhali* trial will shift as participants transition from preconception, to pregnancy, and subsequently to the postpartum follow-up of index children during infancy and early childhood. Consolidating the learnings gleaned from the implementation of the initial preconception and pregnancy phases can therefore provide important feedback for the trial, but also, insights into the mechanisms and challenges specific to preconception and pregnancy interventions, particularly in lower resource settings. Lastly, it can improve understanding around how CHW roles might be further leveraged to strengthen the healthcare sector outside of the trial, and where potential challenges lie. Therefore, the aim of this paper was to describe factors influencing implementation of the preconception and pregnancy phases of the HeLTI *Bukhali* intervention, from the perspective of the HHs delivering the intervention.

## 2. Methods

This study uses qualitative data from focus group discussions (FGDs) collected as part of the trial’s ongoing process evaluation.

### 2.1 Ethical considerations

The Human Research Ethics Committee (Medical) at the University of the Witwatersrand approved the study (M190449). All procedures were carried out according to the Declaration of Helsinki of 1975, revised in 2008, and the HHs gave written informed consent to participate in the study.

### 2.2 Setting and participants

Participants were HHs delivering the *Bukhali* trial intervention in Soweto, South Africa. Supplementary Figure 1 (reproduced with permission (10)) provides an overview of the following four main HH roles within *Bukhali*. Firstly, HHs provide risk screening, through identification, referral and management for obesity, anaemia, hypertension, diabetes, depression, anxiety, and HIV (and pregnancy testing). To assist with risk screening, the HHs use a ‘traffic light’ system, with red indicating high risk requiring referral for further management, orange indicating medium risk, and green indicating low risk, based on established cut-offs per condition. Secondly, HHs provide social support, including emotional support, phase-specific health literacy materials and resources. The third role is to assist with adopting healthier behaviours, using Healthy Conversation Skills and individual-derived goal setting with SMARTER planning (12,13). Lastly, HHs provide and support uptake of multi-micronutrient supplements (MMS) (10). All of the HHs employed by the trial at the time of data collection (n=13) were invited to, and agreed to, participate, between 24 February and 13 March, 2023. The HHs were female, aged 23 to 35 years, with high school degrees and limited additional formal education.

### 2.3 Patient and public involvement

The public was involved in the design, conduct, and dissemination plans of *Bukhali*. Intervention development was guided by formative work conducted with community members (5,8,10,14,15). Engagement with trial stakeholders (e.g. representatives from the South African government, World Health Organisation, and UNICEF) is ongoing, and a participant advisory group has been involved in the qualitative research strategy. The participants of the present study (*Bukhali* HHs) are engaged in regular debrief sessions with their project coordinator and quarterly focus group discussions, through the *Bukhali* process evaluation (8).

### 2.4 Data collection

Three FGDs were conducted between 6 and 13 March 2023 at the trial study site at Chris Hani Baragwanath Hospital in Soweto. The FGDs included 4-5 HHs per group, and lasted 1 hour 35 minutes, 1 hour 58 minutes, and 3 hours 5 minutes. FGDs were facilitated in English by LS and KM, researchers affiliated with the trial but not involved in direct management, evaluation, or day-to-day duties of the HHs. KM was able to translate from participants’ home languages into English, where necessary. A topic guide developed by the co-authors was used, including around HH perceptions of their delivery of the intervention components. The FGDs were recorded and transcribed verbatim by a professional transcriber.

### 2.5 Data analysis

A thematic approach was used to analyse the data, drawing on reflexive thematic analysis as outlined by Braun and Clarke (16), but additionally incorporating more structured elements of a codebook approach to allow for exploration of the pre-determined, process-evaluation driven questions posed by the study (17). Analysis was informed by the UKMRC guidance on process evaluations of complex interventions, which identifies context, implementation, and mechanisms of impact as key components of process evaluation (18). Following familiarisation with the transcripts, a conceptual coding framework was developed by CD, with input from the co-authors. This coding framework identified various factors impacting implementation across the four HH roles. Subsequently, the coding framework was applied to the transcripts, and themes and sub-themes were developed, defined, and refined with input from the co-author team. The resulting revised themes were: interaction with existing public healthcare sector; participant perceptions of health; health literacy and language barriers; participants’ socioeconomic constraints; family, partner, and community views of trial components; and the HH-participant relationship.

## 3. Results

Figure 1 provides a conceptual overview of how the themes relate to the four HH roles.

**Figure 1:**
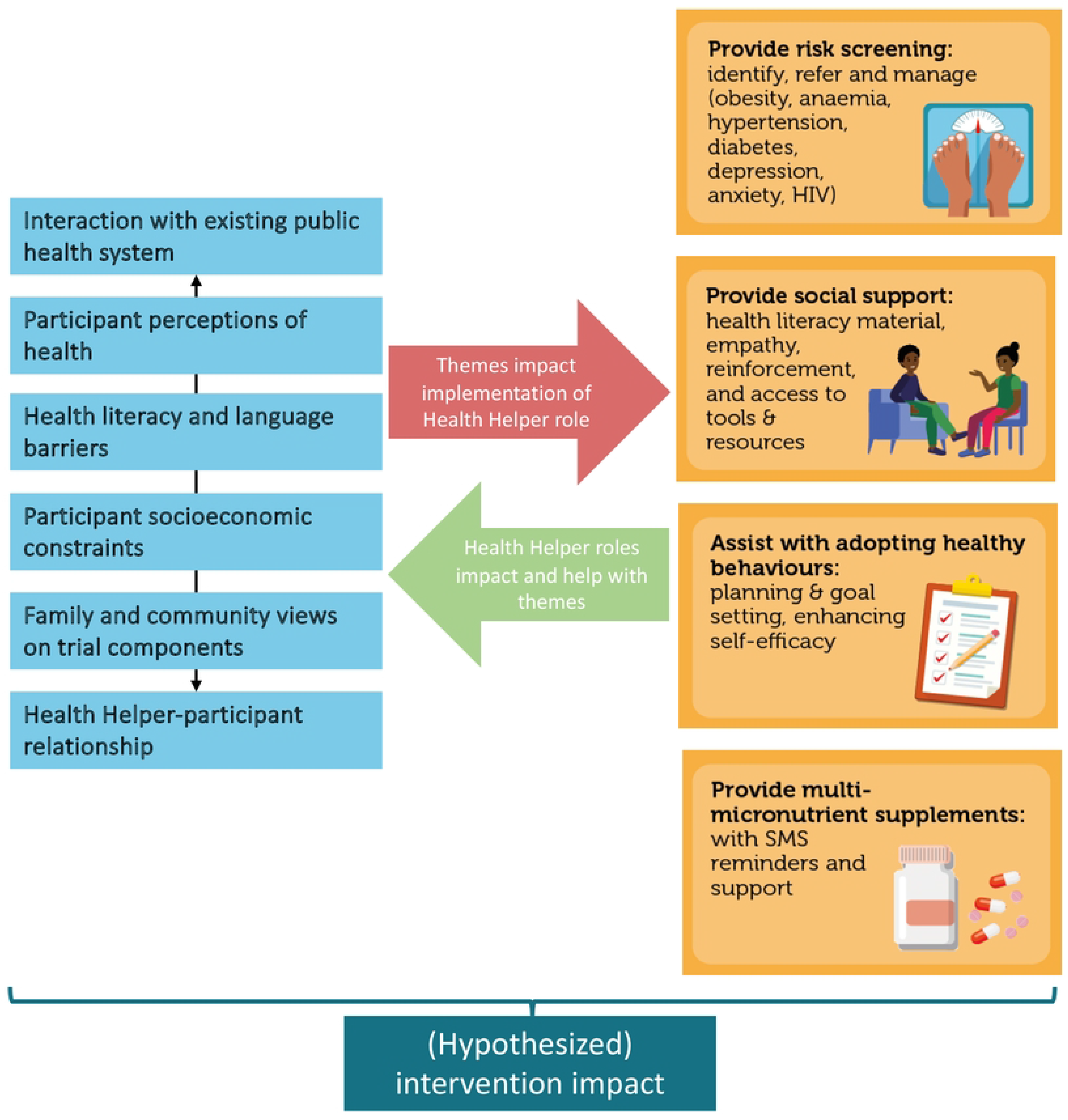
Overview of themes and how these conceptually relate to the four HH roles in the *Bukhali* intervention.

### 3.1 Interaction with existing public healthcare sector: opportunities vs challenges

HHs described how the trial intervention complements existing public healthcare for young women. Although the level of one-on-one care received was different per facility, the main roles that the HHs felt they provided over and above the public health clinics, included provision of necessary information, support and time, and access to resources such as free pregnancy tests and ultrasounds during pregnancy. HH also stated that, in case of health concerns or symptoms, participants often turn to HHs first to ask for advice, including on whether to seek care at the clinic. In this way, the support provided by the HHs seemed to help participants interact with the public healthcare system.

> “I will go back to the support that we give to them. If the participant has a problem we are probably the first people they will ask: ‘I’ve got rash I don’t understand what this rash is’ or, ‘I’m bleeding.’ They won’t go straight to the clinic, so they will always start with us first to find out what we know, what we think, before they go to the actual clinics.” (FGD 2)
>
> “Because at the clinic they don’t really have time to have a one on one session;…Ja, we go as far as explaining the clinic card, you know, and they don’t do that.” (FGD 3)

However, particularly for their role of providing risk screening and referrals, the HHs described challenges of implementing an intervention within the resource-strained health system that it is aiming to complement. Firstly, HHs noted that many participants were reluctant to follow up their referral at public health clinics. Reasons included difficulty getting to the clinic; lack of privacy, particularly for more stigmatised health concerns such as HIV; and poor treatment by clinic staff. Some tools that HHs used to alleviate these challenges included providing trial-based transport to clinics, accompanying their participants to the clinic to ensure they went and were attended to, and emphasising the importance of the referral (Figure 2).

**Figure 2:**
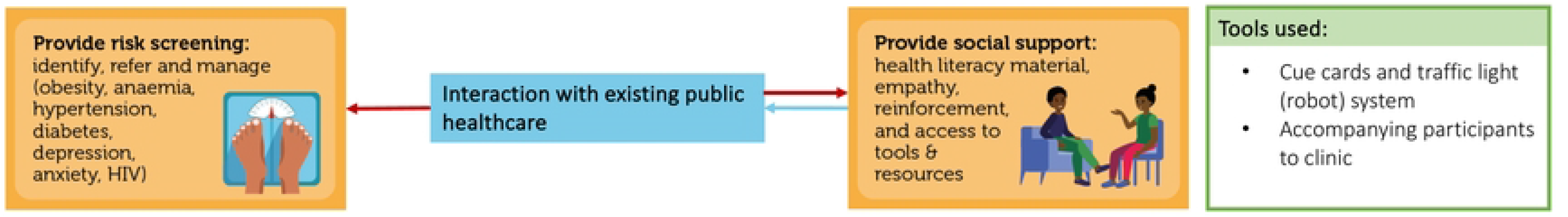
The relationship between the theme “interaction with existing public health system” and the Health Helper roles (orange). Red arrows: negative impact of the theme on role; blue arrows: positive impact between role and theme.

> “Especially if you are going to refer them to the local clinic, they always tell you: ‘The reason why I don’t like going to the clinic is because they shout at us, they don’t like us being there, they ask us why are you here if you are not sick’…so when you refer them they are like ‘yoh no, no, no at the clinic no they’re going to shout at us.’” (FGD 1)
>
> “They are going to try and avoid the clinics by all means.” (FGD 3)

Referrals for mental health concerns such as depression, suicidality, and anxiety were identified as particularly difficult, despite HHs engaging the services of mental health advocacy organisations. HHs expressed that, aside from advocating for their participants to ensure they get help, a trial-based mental health service could be key to filling this gap.

> “I have one incident where we had to go like walk in, me and [Project Coordinator]…so you have to lose your cool in order for you to get something. So they said we must go back to the local clinic so that the local clinic can write a letter for her to come back here and our participant was suicidal so it was hectic, she needed medication like ASAP, ASAP.” (FGD 1)
>
> “And concerning to referral uh my participant like she didn’t get help from [public health service], she had to come back to me and said she don’t want to go back again, she’d rather wait until we have a psychology or a social worker in this study, yes… So I think if we can have our social worker or psychology it would be better.” (FGD 3)

Particularly for health risks such as high blood pressure and low haemoglobin, participants who did go to their clinic often reported back to their HHs that the identified health risk was dismissed at the clinic, with staff insisting no management was necessary. The HHs described that this not only forestalled participant health improvement, but that it also undermined the trust between the HHs and their participants (and thereby their ability to provide support). HHs described feeling powerless in the face of this pattern of inaction by the public clinics: “I was defeated”.

> “And I also think our health facilities are failing us…Because now you send a participant because they are red [high risk on the traffic light system] and they go to the clinic and they test them, with the same results that you got but they tell them that they are okay, they are fine…So it’s a bit of a challenge because now you are saying one thing to them and the clinic is saying something different… They don’t know who to believe and what to do now” (FGD 1)

### 3.2 Participants’ perceptions of health

HHs described a number of ways in which participants’ perceptions of health (and health feedback) influenced implementation of their roles to provide risk screening, assist with healthy behaviours, and provide MMS. In providing health screening feedback, one challenge experienced by HHs was participants not accepting a screening result. For example, participants often did not see themselves as having a high BMI. HHs attributed this in part to cultural norms, with bigger body size representing a cultural ideal, and a lack of awareness of health consequences associated with overweight/obesity. HHs found it helpful to be able to refer to the trial-based dietitian, as an additional expert resource to encourage behaviour change. High blood pressure was reportedly seen by participants as largely an issue of the elderly, and a lack of any notable symptoms made the health risk harder for participants to accept. To help navigate this, HHs described the usefulness of the health screening cue cards which use the traffic light system mentioned earlier, using easy to understand categories of green, orange, and red. HHs also used trial health literacy resources, employed Healthy Conversation Skills and personalised plans for change (SMARTER plans), and emphasised health consequences (sometimes through flawed methods such as scaring participants into action) (Figure 3).

**Figure 3:**
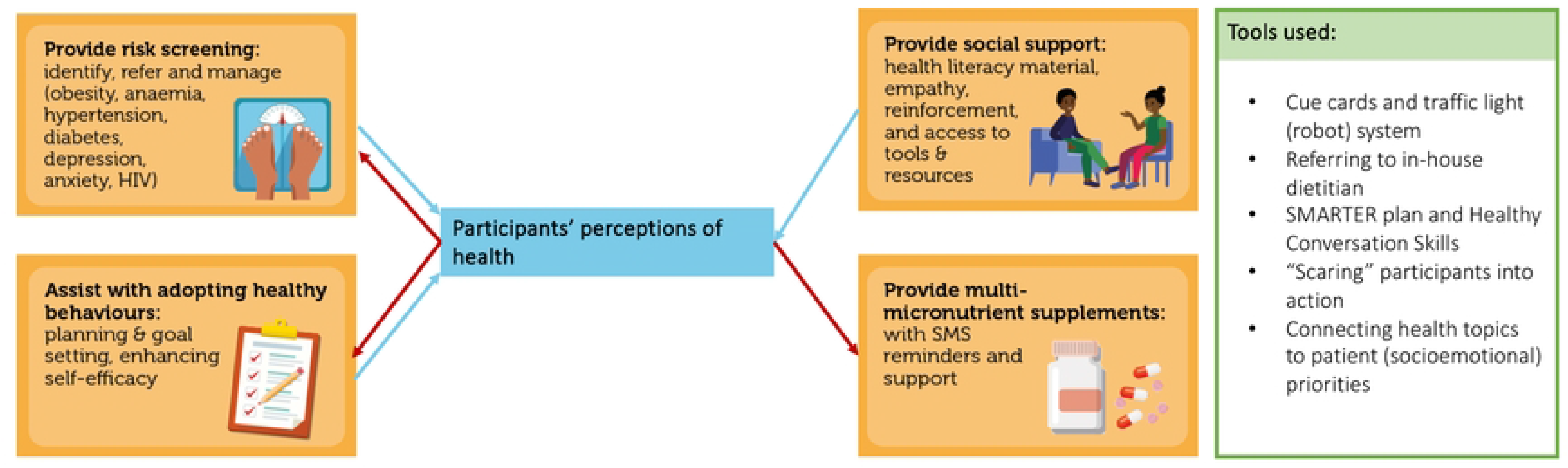
The relationship between the theme “participants perceptions of health” and the Health Helper roles (orange). Red arrows: negative impact of the theme on role; blue arrows: positive impact between role and theme.

> “You find a participant when you let them know like, okay, so your BMI is above 30 which means you are obese, and they say no I’m not, and from there you find that that’s a challenge number one, because now you’re thinking, okay, what am I going to do next, what do I say because this person is saying ‘no, I’m not.’” (FGD 1)
>
> “I’m not feeling sick, I’m okay; why are you saying my BP is this and that; so… ja and some they have this thing that BP uh like high blood is for older people, I’m still young, you can see I’m still, like I can walk and all that, so they don’t believe it when you explain BP to them.” (FGD 3)
>
> “I find that using the robot [traffic light] system is simple… Even the colours, the fact that they see different colours also makes it easier for them to actually understand the results when you give them out to them.” (FGD 2)

In addition, HHs described how participant perceptions of conditions that tend to carry stigma, such as HIV and mental health challenges, impacted their preparedness to openly discuss, accept and act upon these conditions. Participants’ strong emotional response to newly identified positive HIV status was described by the HHs as one of the most challenging aspects of health screening and feedback. In response, HHs used tools such as HIV diagnosis counselling, providing emotional support and space, and accompanying them to the HIV clinic to initiate management.

> “I once had a participant that was the very difficult one, and she was my first participant to test [HIV] positive, yoh, I remember being locked in the office for two hours, she cried.… I allowed her to cry and then once she calmed down we talked about the way forward, I asked her if I could write her [a referral] for [HIV care clinic], but she refused.” (FGD 1)
>
> “The only difficulty that I have is with relating their HIV results. That one is a difficult one because the mood just changes immediately.” (FGD 2)

The HHs noted that participants wanted HHs to discuss socioemotional concerns, prioritising these over physical health topics which seemed to be seen as less relevant to their everyday lives. In such cases, HHs found it helpful to break the ice by discussing and providing support for such socioemotional needs first, and subsequently relating these back to the session’s physical health topics and resources. This aligns with the person-centred Healthy Conversation Skills approach, empowering the participant to identify their own challenges and solutions.

> “They go through a lot of things, different things at home; their backgrounds; so once you touch on the emotions topic they get more comfortable, they open up and they tell you about their lives; I think so, when you want to get a session you need to touch based on the emotions topic a bit, for them to open up.” (FGD 1)

Lastly, HHs found that participants were more receptive of health screening and referrals, health behaviour change, and MMS in the pregnancy phase of the trial, compared to preconception. They attributed this to participants’ concern for their child’s health, which seemed to be received as more tangible and actionable feedback than the often-asymptomatic preconception health risks.

> “It’s [preconception] very tricky. Very. Because you are trying to win them at that stage; you are trying to make sure that they understand the session…And sometimes preconception they tell you sometimes that I don’t feel anything, I feel fine.” (FGD 3)
>
> “At pregnancy and infancy they are scared something will happen to the baby… And then they will immediately go to the clinic. Even if they shout at them, they will go.” (FGD 1)

### 3.3 Health literacy gaps and language barrier

Participant health literacy impacted how HHs provided risk screening (feedback) and assisted with adopting healthy behaviours. Depending on participants’ prior knowledge and education level, reported topics with poorer health literacy amongst participants included NCDs, mental health, sexually transmitted illnesses, and healthy diet and sleeping behaviours. These gaps required HHs to tailor the sessions according to participants’ needs. For example, HHs dedicated effort and time to finding creative ways to explain content and avoid medical jargon, simplifying key messages per session, repeating key pieces of information between sessions (Figure 4).

**Figure 4:**
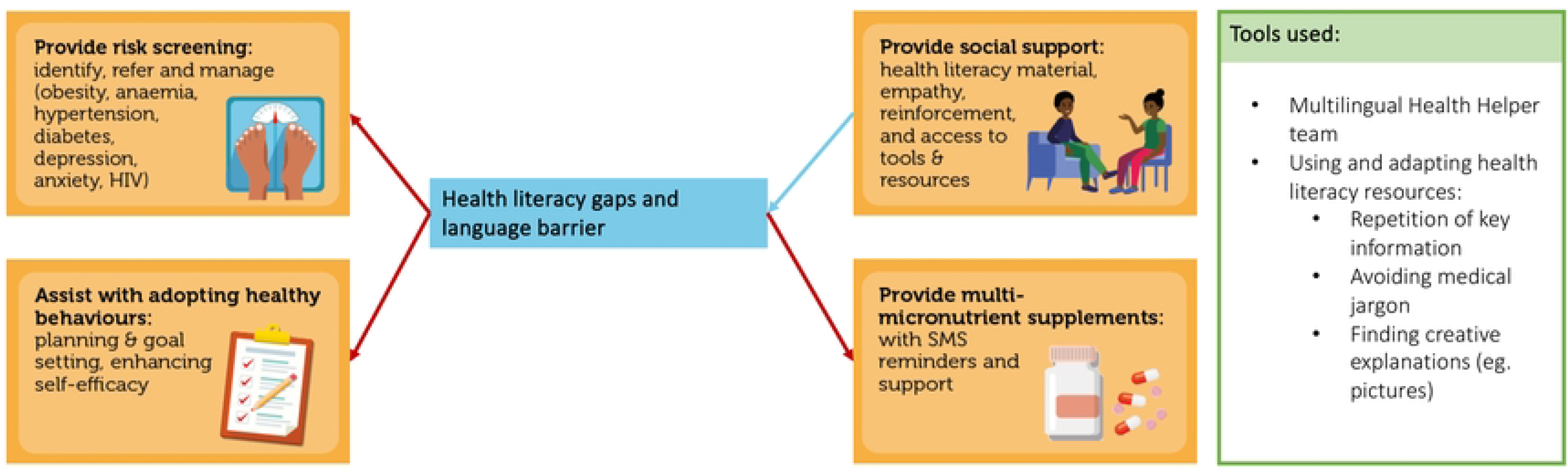
The relationship between the theme “health literacy gap and language barrier” and the Health Helper roles (orange). Red arrows: negative impact of the theme on role; blue arrows: positive impact between role and theme.

> “You give them the results when they come for their 6 months and when they come back again and ask them, “Do you still remember your results? Do you still understand?” They don’t know anything. So you need to go back again and recap everything.” (FGD 2)
>
> “We usually make them [blood pressure] sounds because it’s easier. Imagine telling someone the systolic is like this, they don’t know what systolic is, so we have to be like (knocks on table)… And then they’re like oh okay, then they get to understand what the word is; so I wouldn’t say that there’s resistance, I’d just say that it’s a bit tricky changing those words into something they will understand. You have to be creative.” (FGD 3)

Another identified challenge was that some participants experience a language barrier and are not able to read the trial’s English resource books, making it difficult for them to engage with the trial resources. In these cases, HHs needed to spend time explaining and verbally translating the resources during their session, enlist the help of a colleague who speaks the participant’s home language if necessary, and use pictures to explain the content. This was anticipated in the planning of the intervention, but was considered more feasible and preferrable to translating all the materials into the multiple possible languages (South Africa has 11 official languages), when the written version of participants’ home language may not necessarily be preferred.

> “I think with the language barrier you can switch with one of the HHs who actually understands that language for them to assist you with that regard…You need to try by all means… Maybe opening Google as well showing them pictures from there as well, that also helps. Making examples, maybe using yourself as an example.” (FGD 2)

### 3.4 Participants’ socioeconomic circumstances

Participants’ socioeconomic realities, especially food insecurity and unemployment, were found to impact how participants received and were able to act upon the intervention content. Specifically, participants could be prevented from adopting healthier behaviours (dietary and sleep behaviours), following through on referrals (due to lack of transport to healthcare clinics), and taking MMS (Figure 5).

**Figure 5:**
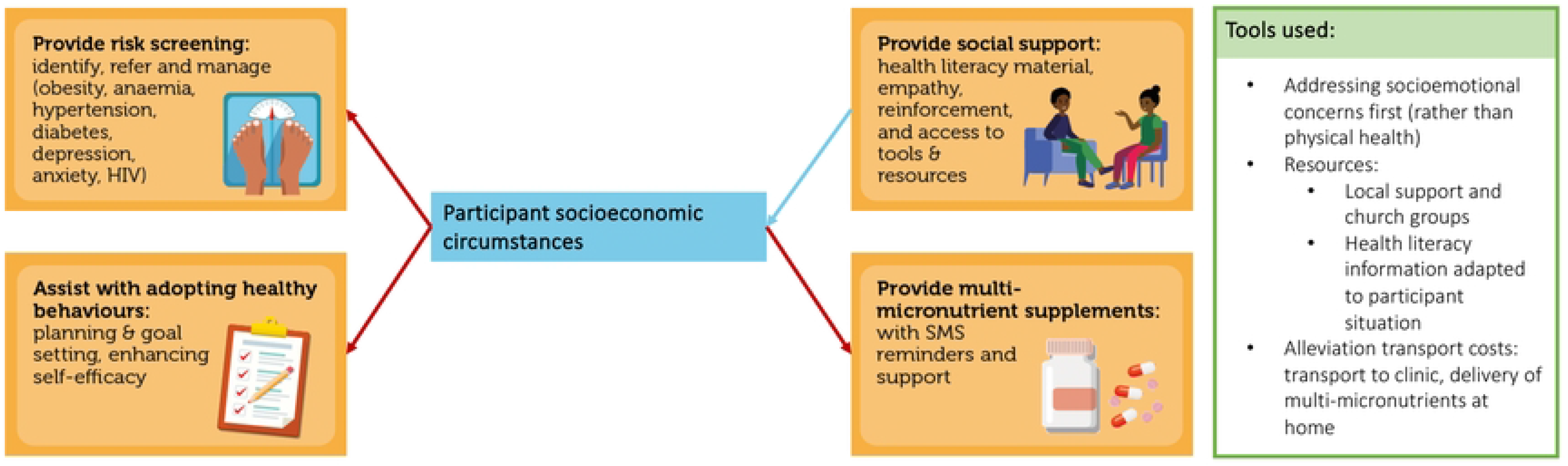
The relationship between the theme “participants’ socioeconomic circumstances” and the Health Helper roles (orange). Red arrows: negative impact of the theme on role; blue arrows: positive impact between role and theme.

> “It is always a challenge with unemployment. Not having enough food to eat, yeah…It is because you eat what’s in the house. You eat what’s cooked. You can’t tell them that “I don’t want this, I want that…” So, it is a very tricky one when it comes to diet. It is very tricky.” (FGD 2)
>
> “Most of them they are sleeping because they are unemployed.” (FGD 1)

In such situations, HHs informed participants of local church groups who can assist, but also described occasionally going above and beyond to help participants in a personal capacity. Lastly, socioeconomic issues, including substance abuse, were described as contributing to challenges tracing participants, as participants in these circumstances were more likely to move between addresses.

> “Like, we are expected to find them, they change locations… it becomes a challenge when you get there, their parents don’t know where the girl is, they started using drugs, they live in a drug house somewhere, now it becomes unsafe for us as well to go and track them.” (FGD 3)

### 3.5 Family, partner, and community views of trial components

The HHs identified family, partner, and community views of trial components as influential to whether participants were able to enact the intervention (Figure 6). This included negative views on MMSs and lack of support for adopting healthy behaviours, particularly in terms of diet. The participant’s living situation influenced the way family or her partner impacted implementation, highlighting the importance of integrating intervention aspects such as supplementation and dietary behaviour within families and communities.

**Figure 6:**
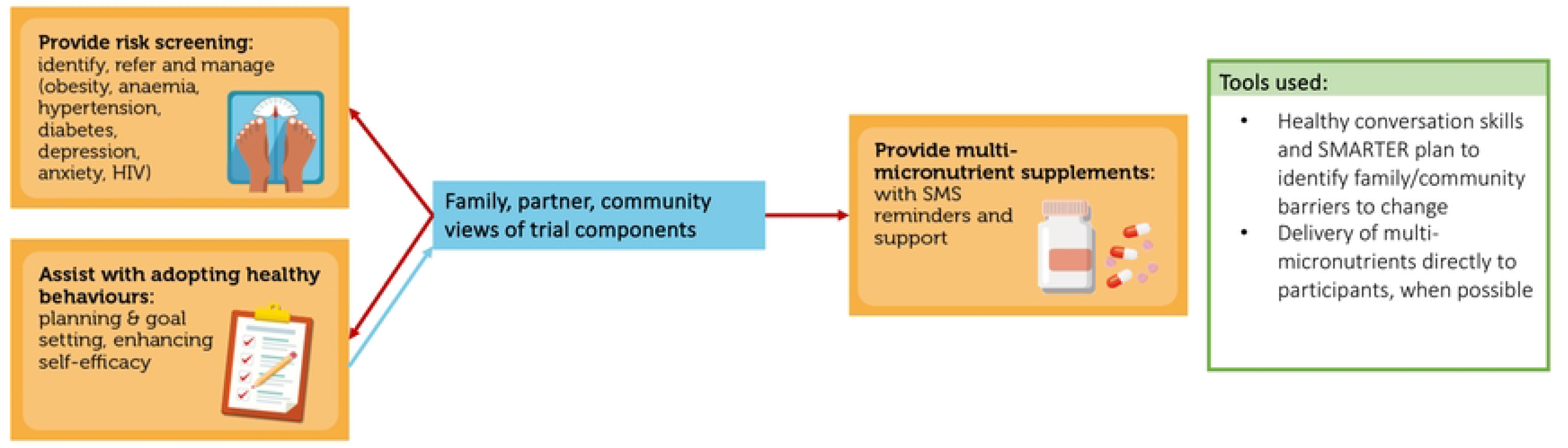
The relationship between the theme “family, partner, and community views of trial components” and the Health Helper roles (orange). Red arrows: negative impact of the theme on role; blue arrows: positive impact between role and theme.

> “‘Cause of the beliefs at home, uh they don’t believe in using any form of supplements.” (FGD 1)
>
> “I think their backgrounds, their backgrounds; because sometimes it’s hard to change a behaviour if you are not being supported; if at home that is what they eat and you now come all of a sudden and you are like let’s change this, then they are all going to ask you why must we change it because of you, and now it’s going to seem like you are the bad person.” (FGD 1).

Community stigma around sickness, and particularly around HIV and taking medication (which supplements were perceived as), also impacted the degree to which HHs were able to effectively assist with behaviour change and risk management (see also section 3.2). On the other hand, MMS were also sometimes shared between family members, preventing optimal intake by participants.

> “Another thing is the stigma; if we go to the locations and we’re carrying these small packages and then we’re like I’m going to deliver iron supplements then they’ll be prying like what is iron supplements, how do you take them, Why are they so important; and then they’ll be like isn’t she sick?… sick like as in HIV or something.” (FGD 1)
>
> “And then the 80% has challenges when it comes to supplements, ‘no I can’t take them because of Gogo [grandmother] took them’, ‘I can’t take them because of at home they’ll think I’m sick.’” (FGD 2)

### 3.6 Health Helper-participant relationship

A relationship of trust between the HH and participant was seen as critical to providing risk screening (making it more likely that participants go to the clinic when advised to do so), assisting with healthy behaviours, providing social support, and supporting the uptake of MMS [Figure 7]. HHs commented on the challenge of switching between a supporting, trusted role and their more practical tasks, such as completing session content and data capturing. In addition, the bond of trust occasionally created a challenge when another HH needed to assist a participant. This could be navigated if the HH vouches for the trustworthiness of their colleague stepping in.

**Figure 7:**
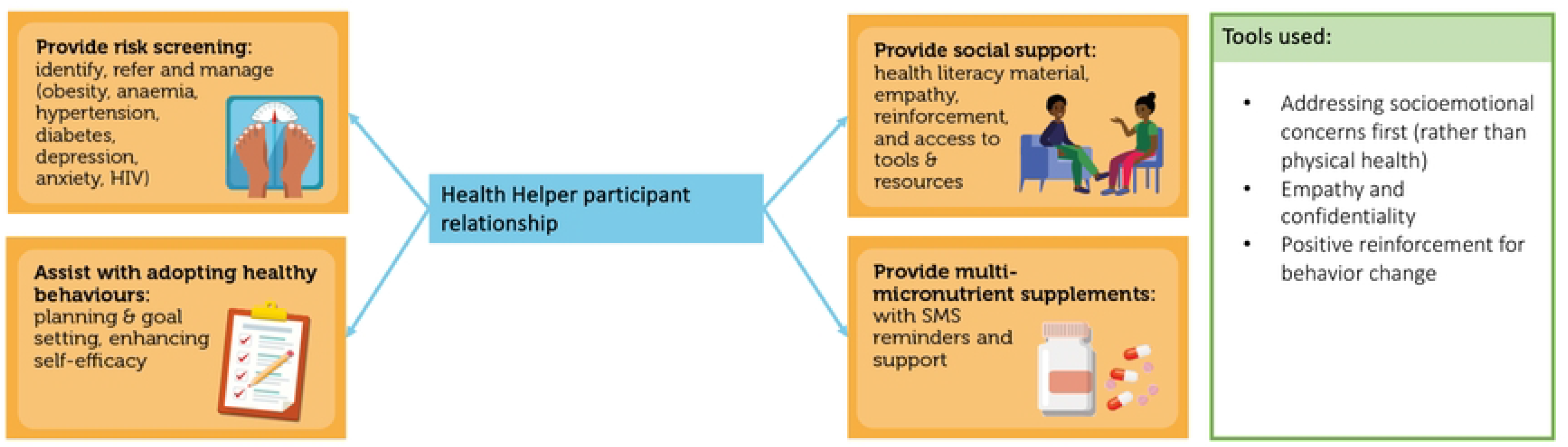
The relationship between the theme “Health Helper-participant relationship” and the Health Helper roles (orange). Blue arrows: positive impact between role and theme.

> “As [HH] said that we are the first that they come to, so, I think it is because of we give them an opportunity to build trust together. Because she knows that the moment she leaves, whatever she said to [HH], it is between her and [HH]. So it is private, even when she gets home nobody is going to know about it.” (FGD 3)

## 4. Discussion

This paper reports on HHs’ perspectives of the implementation of the initial preconception and pregnancy phases of *Bukhali*. The results indicate that the HHs are largely able to implement their roles as intended, despite facing context-related challenges to implementation of the intervention. These challenges stemmed from barriers within the public health system, participant perceptions of health, limited participant health literacy, participant socioeconomic constraints, and family and community views of trial components. The HHs also highlighted tools that they use to navigate these challenges, and emphasised previously documented findings around the importance of the HH-participant relationship is a mechanism of implementation, which can help to alleviate some of the identified implementation challenges (9). These findings provide support for a number of implementation adaptations specific to the *Bukhali* intervention, which fall into three more broadly applicable strategic areas for optimising CHW-delivered preconception and pregnancy interventions: navigating and bridging health care systems, adaptability to individual patient needs, and navigating stigma around disease.

The strategic potential of HHs to support participants in navigating the public healthcare system aligns with a well-described role for CHWs to improve continuity and bridge gaps between healthcare sectors (6,19,20). From our results, this even included HHs’ being participants’ first call in case of a health emergency, an unintended form of task shifting towards HHs that requires adequate training to empower HHs to react appropriately in such situations (21).

Additionally, the lack of management and treatment by clinics in response to some participants’ referrals, particularly for hypertension, was identified as a challenge. Aside from possibly impacting trial outcomes, this finding is concerning for participants’ health, as preconception hypertension presents a risk for future cardiovascular disease in mothers (22–24), and for adverse outcomes in a potential pregnancy (25–27). The reported undermanagement may reflect a lack of current preconception care within the South African public healthcare system (28–30). In response to this finding, the *Bukhali* intervention team plans to implement more extensive follow-up of these high-risk, unmanaged cases. This will be done through guideline-based confirmatory testing of hypertension on three separate occasions (NDOH national user guideline), followed by a more hands-on, directive referral to a point of (if necessary, tertiary) care. This will help participants to navigate the referral pathway with more support from the trial and their HH, help prevent their hypertension from going untreated, and hopefully provide a clearer understanding of why the clinics are not be providing management in these cases. Within our context, this challenge also illustrates how increased health screening during preconception and pregnancy introduces an increased caseload within an under-resourced healthcare system, requiring policy-level investments into primary prevention. This could include, for example, solutions such as private-public healthcare partnerships (31).

Within an urban setting that is heterogenous in terms of participants’ education, food security, and cultural and linguistic backgrounds, many of the tools described by the HHs reflect the need to adapt intervention delivery to cater for individual participant needs and/or circumstances. Examples from this analysis include switching between languages, finding creative ways to explain the resources across levels of education, and helping participants find individualised solutions to socioeconomic barriers to participation. Although this kind of adaptability can have implications for fidelity to the study protocol, an adaptable, pragmatic approach to a CHW-delivered intervention improves the applicability and feasibility of the implementation (10,32). For *Bukhali*, our results highlighted the importance of ongoing training of the HHs, with emphasis on how to adapt sessions and explanations in multiple ways, using pictures, drawings, and different languages and avoiding adverse tactics such as ‘scaring participants into action‘. In the broader context of CHW programmes across South Africa, which requires coverage across demographics, cultural variations, and rural versus urban settings, programmes will likely benefit from an emphasis on adaptability on CHW programme level (2), and on the individual CHW-patient level (9). Achieving this may require additional investments in CHW training, support, and resources.

Lastly, the themes of ‘participant perceptions of health’ and ‘family and community views on trial components’ illustrated that HHs needed to navigate stigma around sickness and disease. In South Africa, the widespread impact of the HIV epidemic and associated stigma provide important context to stigma and views around (chronic) disease and taking medication, more generally (33–37). For example, a correlation between HIV stigma and stigma around COVID-19, was recently documented amongst people living with HIV (38). For *Bukhali,* HIV-related stigma was found to impact participants’ reluctance to test for HIV, their denial around HIV diagnoses, and their fear of judgement when taking MMS. In this context, the design of preconception and pregnancy interventions should acknowledging the impact of health-related stigma, and its intersection with stigma related to other factors such as race, gender, and class (39,40), on implementation practices. For *Bukhali*, this has signalled the need for ongoing community engagement and education efforts with a focus on de-stigmatisation of health conditions, in parallel to the trial implementation.

The results of this study emphasise the importance of reporting process evaluation outcomes at an early point in the trial. Particularly in a multi-phase trial of such long duration, this allows for the reporting of phase-specific findings, and for suggested improvements to be applied to the ongoing trial. One limitation of this approach is that it provides a snapshot of current perspectives on the implementation of *Bukhali*, rather than an overview representative of the whole trial and each of its phases. However, as part of the process evaluation we will continue examining the implementation longitudinally and across all phases. Secondly, since the HHs were employed by the trial, power dynamics between the researchers/interviewers and HHs may have influenced their responses in the FGDs.

However, in order to minimise any such effect, the FGDs were facilitated by researchers who were not directly involved in the management or day-to-day work of the HHs.

## 5. Conclusions

In conclusion, this qualitative evaluation of HH perspectives of an ongoing multi-phase randomised controlled trial identified three main strategic areas for optimising CHW-delivered interventions in our setting, namely navigating the healthcare system, adapting to individual participant needs and circumstances, and navigating health-related stigma.

Within all three of these strategies, the HH/CHW-participant relationship was found to be a pivotal mechanism for the interventions’ impact. These findings provide insights and recommendations for the next phases of the *Bukhali* intervention, for other CHW-delivered preconception and pregnancy trials, and for the strengthening of ‘real-world’ CHW roles in settings with similar implementation challenges.

## Data Availability

The authors do not have permission to share the data for this study, due to ethical concerns from the relevant Human Resource Ethics Committee about sharing qualitative interview data outside of the research team.

## Acknowledgements

We thank the participants and staff of the *Bukhali* trial for their contribution to this study.

## Supporting information captions

Supplementary Figure 1 Overview of the four Health Helper roles within *Bukhali*. Reproduced with permission (10).

